# Safety and immunogenicity of an inactivated SARS-CoV-2 vaccine, KD-414, in healthy adult and elderly subjects: a randomized, double-blind, placebo-controlled, phase 1/2 clinical study in Japan

**DOI:** 10.1101/2022.06.28.22276794

**Authors:** Mitsuyoshi Tanishima, Kayo Ibaraki, Keishi Kido, Shun Nakayama, Kohei Ata, Hideki Nakamura, Yasuhiko Shinmura, Masafumi Endo, Kengo Sonoda, Kohji Ueda, Yoshiaki Oda

**Author notes:** **CORRESPONDANCE** Mitsuyoshi Tanishima, KM Biologics Co., Ltd (KM Biologics), 869-1298 Kumamoto, Japan.

## Abstract

**Background:** In the current protracted COVID-19 pandemic, SARS-CoV-2 vaccines that have the ability to be used safely and to prevent onset or severe disease are still highly needed. A Phase 1/2 study was conducted in healthy adults and the elderly in Japan to evaluate the immunogenicity, safety, and tolerability of an inactivated whole-virus vaccine (KD-414) that is under development.

**Methods:** In this double-blind, randomized, placebo-controlled, Phase 1/2 study, adults aged 20 to 64 years and elderly participants aged 65 years or older without a history of COVID-19 were randomly allocated to the following groups: the L group (2.5 μg/dose), M group (5 μg/dose), or H group (10 μg/dose) with KD-414, or the placebo group (2:2:2:1). The participants received KD-414 or the placebo intramuscularly twice at intervals of 28 days. To determine the go-forward dose, safety after the first dosing and neutralizing antibody titers against SARS-CoV-2 at 28 days after the second dosing were evaluated for each group. Additionally, after unblinding, participants in the H group received a third dose of KD-414 (H) approximately 6 months after the second dosing for an exploratory evaluation of the safety and neutralizing antibody titers to be conducted.

**Results:** A total of 210 participants were enrolled: 105 adults aged 20 to 64 years, and 105 elderly participants aged 65 years or older. Of these participants, 105 adults and 104 elderly participants completed the second dosing, and 28 adults and 31 elderly participants in the H group received a third dose of KD-414 (H). The incidence of adverse reactions from the first dosing to 28 days after the second dosing was 19 of 30 (63.3%), 22 of 31 (71.0%), 22 of 29 (75.9%), and six of 15 (40.0%) for adults, and 14 of 30 (46.7%), 14 of 29 (48.3%), 15 of 31 (48.4%), and six of 15 (40.0%) for elderly participants in the L, M, H, and placebo groups, respectively. No differences in incidence were shown among the KD-414 groups. The most common adverse reaction was injection site pain. Fever that resolved the following day was observed in only 1 adult in the H group after the second dosing; this was a sole Grade 3 or higher adverse reaction. For immunogenicity, the neutralizing antibody seroconversion rate (95% confidence intervals [CI]) against SARS-CoV-2 (vaccine strain) 28 days after the second dosing was 36.7% (19.9-56.1), 38.7% (21.8-57.8), and 72.4% (52.8-87.3) in adults, and 33.3% (17.3-52.8), 31.0% (15.3-50.8), and 45.2% (27.3-64.0) in elderly participants in the L, M, and H groups, respectively, showing a dose response by KD-414. The stratified analysis by age-range for the H group, which observed the highest immunogenicity, also showed an age dependency in the neutralizing antibody responses. Based on these results up to the second dosing, the H (10 μg/dose) dosage was determined as the recommended dosage for further clinical development of KD-414. In addition, there was no particular difference between the incidence of adverse reactions after the third dosing and that after the second dosing with KD-414 (H) in participants. Moreover, the geometric mean neutralizing antibody titers (GMTs) against SARS-CoV-2 (vaccine strain) 28 days after the third dosing were 2-fold higher than those at 28 days after the second dosing, and the GMTs 13 weeks after the third dosing were 3-fold higher than those at 13 weeks after the second dosing. The stratified analysis by age-range of Pseudovirus SARS-CoV-2 (D614) spike protein neutralizing antibody titers showed 100.0% neutralizing antibody seroconversion rate and high neutralizing antibody titers in participants aged ≤ 40 years.

**Conclusion:** KD-414 was well tolerated in healthy adults and the elderly at all doses evaluated. In view of the dose-response and age-dependency of the immunogenicity of KD-414 (H) (10 μg/dose), it is expected to induce high neutralizing antibody titers, particularly in the age range of 20 to 40 years. A Phase 2/3 study (Japan Registry of Clinical Trials [jRCT] 2071210081), a Phase 3 study (jRCT 2031210679), and a Phase 2/3 study in pediatric participants aged 6 months to 17 years (jRCT 2031220032) using KD-414 (H) are ongoing.

## Introduction

COVID-19 is an infectious disease caused by severe acute respiratory syndrome coronavirus 2 (SARS-CoV-2), which was identified in December 2019 in Wuhan, Hubei Province, China. Thereafter, a Public Health Emergency of International Concern (PHEIC) was declared by the World Health Organization (WHO) on January 30, 2020 and a global pandemic was declared on March 11, 2020^1^. As of May 22, 2022, 522 million cases of SARS-CoV-2 infection have been reported, resulting in 6 million deaths, so vaccine development against COVID-19 remains a significant global challenge^2^. At present, vaccines against COVID-19, including inactivated vaccines, DNA vaccines and mRNA vaccines, viral vector vaccines, recombinant protein vaccines, etc. have been developed in the United States, the United Kingdom, China, and other countries around the world. In Japan, four products of mRNA vaccines, a viral vector vaccine, and a recombinant protein vaccine have been approved as of May 2022^3-6^. However, most of them are imported and there are concerns regarding adverse reactions associated with new modality, such as myocarditis and thrombosis with thrombocytopenia syndrome^7-10^. Because of that, booster shots (a third dose) and primary vaccination in children who require higher safety are not progressing. It is important to secure sufficient amounts of vaccines against SARS-CoV-2 in Japan for future emergencies and, similar to seasonal influenza vaccines, these vaccines will be continuously required in the future. Thus, domestic vaccines with high safety are needed.

KD-414 is a purified, inactivated, whole-virus, SARS-CoV-2 vaccine developed by KM Biologics Co., Ltd. (Kumamoto Japan). Inactivated vaccines have been used extensively over decades for multiple infectious diseases, and the safety of these vaccines has been established.

The results of non-clinical studies showed no toxicological changes related to KD-414 in repeat-dose toxicity studies, and induction of neutralizing antibodies and protection against the SARS-CoV-2 challenge in efficacy studies.

Based on these results, we conducted a Phase 1/2 study to investigate the safety and immunogenicity of two or three doses of KD-414 in adults aged 20 to 64 years and elderly participants aged 65 years or older.

## Materials and Methods

### Study design, participants

This study was conducted at two study sites in Japan as a double-blind, randomized, parallel-group, placebo-controlled study. Participants were healthy adults aged 20 to 64 years and healthy elderly participants aged 65 years or older, with the target sample size being 210 participants in total (105 adults and 105 elderly). Each participant was allocated to one of four groups to receive either one of three formulations of KD-414 or a vehicle, and was vaccinated with two intramuscular doses (0.5 mL per dose) at an interval of 28 days. In addition, some participants received a third dose approximately 6 months after the second dosing.

Hakata Clinic Institutional Review Board gave ethical approval for this study. Individuals with a history of COVID-19 or vaccination against COVID-19 were excluded. If SARS-CoV-2 infection was identified during the study, the participant was withdrawn from the study. Written informed consent was obtained from all participants before the start of screening. The study was conducted in accordance with the ethical principles in the Declaration of Helsinki, Good Clinical Practice (GCP) of Japan, and other relevant laws and regulations. The study is registered in jRCT (jRCT 2071200106).

### Vaccine

KD-414 is a whole-virus inactivated SARS-CoV-2 vaccine manufactured using a virus strain (SARS-CoV-2/UT-HPCo-038/Human/2020/Tokyo) isolated from the sera of a Japanese patient with SARS-CoV-2 infection at the Institute of Medical Science, University of Tokyo. KD-414 is a vaccine in which a virus proliferated in Vero cells (African green monkey kidney cell-derived cell line) was inactivated and purified, and formulated with aluminum hydroxide as an adjuvant. In this study, three dose formulations consisting of different amounts of total proteins, Formulation H (10 μg/dose) manufactured under compliance with Good Manufacturing Practice (GMP), the 2-fold diluted Formulation M (5 μg/dose), and the 4-fold diluted Formulation L (2.5 μg/dose) were used. In addition, a mixture of equal volumes of adjuvant and physiological saline (vehicle) was used as a placebo.

### Randomization and masking

Adults and elderly participants were randomized at a 2:2:2:1 (L group: M group: H group: placebo group) ratio using a stratified permuted block randomization method at each study site. Allocation factors were sex and age for adults and sex for elderly participants to minimize these biases. The study was conducted in a double-blind manner, and all participants, principal investigators, and staff remained blinded until the time of unblinding.

### Trial procedures

After no serious adverse reactions or Grade ≥3 adverse reactions were confirmed up to 6 days after the first dosing in the sentinel group (approximately 15 to 20 adults), vaccination progressed for the remaining adults and elderly participants. Each participant was required to record injection-site and systemic adverse events and body temperature up to 6 days after each vaccination. In addition, participants received the second dose 4 weeks after the first dosing and underwent follow-up examinations 4 weeks later. After the follow-up examinations were completed, the participants entered a 1-year follow-up period.

Blood samples were collected to measure neutralizing GMTs before each vaccination, 4 weeks after each vaccination; and 14 days and 13, 26, 52 weeks after the second vaccination. In addition, blood sampling was also performed 7 days after the first and second vaccinations for some of the participants (approximately 20 each of adults and elderly participants) except for the sentinel group, in order to confirm the Th1/Th2 balance (ratio of IFN-γ/CD4+ cell count to IL-4/CD4+ cell count) and Th1/Th2 cytokine production.

Because KD-414 (H) was selected as the recommended dosage after unblinding, participants in the placebo, L, and M groups had completed this study and were recommended to participate in the public vaccination program.

Participants in the H group received a third dose of KD-414 (H) at least 26 weeks after the second dosing, and they are in ongoing follow-up after the third dosing. Participants who did not wish to receive the third dose are under ongoing follow-up after the second dosing.

Solicited injection-site adverse events were defined as erythema, swelling, induration, and pain that occurred up to 6 days after each vaccination and were considered to be causally related to the investigational product. Fever, headache, fatigue, nausea, and muscle pain that occurred up to 6 days after vaccination were defined as solicited systemic adverse events. All events that were not classified as solicited adverse events were recorded as unsolicited adverse events. The reported adverse events were graded according to the criteria from the United States Food and Drug Administration (FDA)^11^, however, for erythema/redness, induration/swelling, and fever, those less than Grade 1 were evaluated as Grade 0.

The primary immunogenic endpoint was the seroconversion rate to neutralizing antibodies against SARS-CoV-2 (D614G) 28 days after the second dosing in each group.

Neutralizing antibody seroconversion was defined as a rise in neutralizing antibody titers to 1:10 (for live SARS-Cov-2 (D614G)) or 80 (for pseudovirus) from seronegative at baseline (before the first dosing) or a four-fold titer increase if the participant was seropositive at baseline. The positive cut-off of neutralizing antibody to live SARS-Cov-2 (D614G) was 1:5, and neutralizing antibody to pseudovirus was 40.

### Neutralizing antibody titration assay

#### Live SARS-CoV-2 (D614G) neutralizing antibody titers

Live SARS-CoV-2 (D614G) neutralizing antibody titers were measured by KM Biologics Co., Ltd. using the 50% Tissue Culture Infectious Dose method. The assay was validated at KM Biologics. Briefly, a mixture of viral solution (vaccine strain: SARS-CoV-2/UT-HPCo-038/Human/2020/Tokyo [D614G]) and serially diluted serum samples was added to VeroE6/TMPRSS2 cells and incubated until cytopathogenic effects were observed. After incubation, the cells were inactivated with formalin. After staining with naphthol blue black, the cells were lysed with 0.1 N NaOH and the absorbance at 630 nm was measured using a plate reader. The cutoff value (([mean absorbance of cell control wells] + [mean absorbance of virus control wells]) × 0.5) was calculated, and wells below the cutoff value were judged as virus-positive wells. The neutralizing antibody titer was the maximum dilution factor at which virus-negative wells with absorbance higher than the cutoff value were confirmed.

#### Pseudovirus SARS-CoV-2 (D614) spike protein neutralizing antibody titers

The pseudovirus neutralizing antibody assay^12^ was performed at LabCorp Drug Development. The pseudovirus neutralizing antibody assay with a lentivirus-based pseudovirus particle expressing the SARS-CoV-2 (D614) spike protein was validated at Monogram Biosciences and then transferred to LabCorp Drug Development. Briefly the mixed pseudoviruses and serially diluted serum samples were incubated in HEK293 cells expressing the ACE2 receptor. After incubation, the luciferase produced was measured to determine the pseudovirus neutralizing antibody titers (50% inhibition dose [ID50]) using the following calculation formula. The LLOQ for pseudovirus neutralizing antibodies is 40 (ID50).

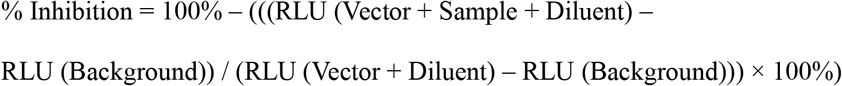

#### Th1/Th2 balance

Whole blood was nonspecifically stimulated with phorbol 12-myristate 13 acetate, and the cytokines for which production was stimulated were accumulated in cells treated with brefeldin-A. After sorting for CD4+ cells using an anti-CD4 antibody, CD4+ cells were counted by flow cytometry after intracellular staining using monoclonal antibodies against IFN-γ and IL-4. The ratio of positive Th1 cells (IFN-γ) and Th2 cells (IL-4) among the CD4+ cells was defined as the value for Th1/Th2 balance.

#### Th1/Th2 cytokine assay

Participant serum samples were analyzed with the Bio-Plex Pro Human Cytokine Th1/Th2 9-Plex Panel (Bio-Rad, #M5000005L3) in accordance with the assay instructions provided. This panel includes 9 cytokines and chemokine cell signaling molecules (IL-2, IL-4, IL-5, IL-10, IL-12 (p70), IL-13, GM-CSF, IFN-γ, TNF-α).

### Statistical analyses

Given the small number of participants in each group, the study was not powered for formal statistical comparisons between groups or time points. The sample size was set based on the minimum sample size required to evaluate safety and immunogenicity. Thus, adjustments for Type I error multiplicity were also not performed. In addition, in all analyses, missing data were not imputed. Statistical analyses were performed separately in adults (20-64 years) and the elderly (≥65 years).

The safety analysis was performed in the population vaccinated with KD-414 with evaluable safety data. The number of participants with solicited injection-site reactions, solicited systemic adverse reactions, and unsolicited adverse reactions and the incidence of each were calculated. In addition, an exploratory evaluation of Th1/Th2 balance was performed in a portion of participants and descriptive statistics for the Th1/Th2 (CD4) cell ratio were calculated.

Immunogenicity was analyzed in the full analysis set (FAS) and the per protocol set (PPS), with the FAS serving as the primary population. The FAS was a population of participants who received KD-414 excluding participants without neutralizing antibody titer data. In the PPS, major protocol deviations (related to inclusion criteria, dosage and administration, concomitant medications, and assessment points) were excluded from the FAS. The seroconversion rate and Clopper-Pearson 95% CI for neutralizing antibody titers 28 days after the second vaccination were calculated as the primary endpoint, and the geometric mean titer (GMT) and 95% CI were calculated as secondary endpoints. These analyses were performed at follow-up time points in the same manner. In addition, subgroup analyses were performed in the H group by age stratification by 10 years, and the pseudovirus SARS-CoV-2 (D614) ID50 GMTs were converted to the WHO international standard units (IU/mL) using the conversion factor “0.1458 (for geometric mean)” determined with the WHO International Standard (20/136) informed by LabCorp Drug Development^12^.

For other background factors, descriptive statistics were calculated for continuous data, and frequencies and percentages were calculated for categorical data.

SAS Release 9.4 (SAS Institute inc., Cary, NC, USA) was used for the statistical analyses described above.

## Results

### Population

A total of 210 participants were enrolled in this study, and 105 adults (L group: 30, M group: 31, H group: 29, placebo group: 15) and 105 elderly participants (L group: 30, M group: 29, H group: 31, placebo group: 15) received at least one dose of vaccine or a vehicle. All participants completed immunogenic evaluation at Day 28 except for one (elderly, placebo group) who was withdrawn after the first dosing. 105 adults (L group: 30, M group: 31, H group: 29, and placebo group: 15) and 104 elderly participants (L group: 30, M group: 29, H group: 31, and placebo group: 14) were eligible for the immunogenic evaluation (FAS and PPS). All participants were included in the safety analyses. Twenty-eight adults and 31 elderly participants received the third dose of KD-414 (H) and were eligible for the safety and immunogenic evaluation after the third dosing (Figure 1).

**Figure1:**
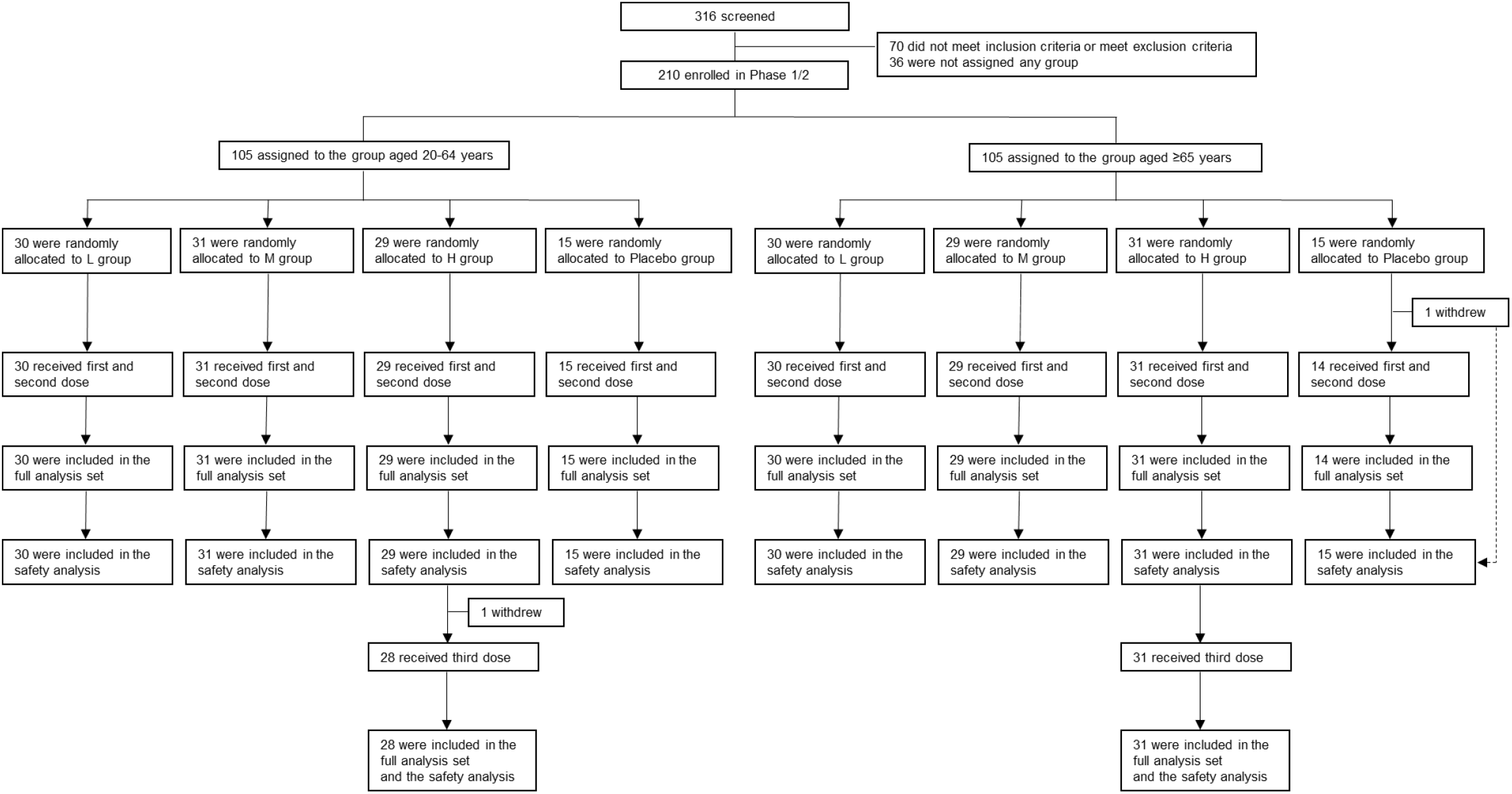
Trial profile

A total of 22 adults (L group: 5, M group: 7, H group: 9, placebo group: 1) and 22 elderly participants (L group: 6, M group: 5, H group: 8, placebo group: 3) were included in the immune response analysis set to conduct exploratory evaluations of the Th1/Th2 balance and cytokine production.

The mean age in each group was 42.8 to 49.6 years in adults. Baseline demographic characteristics of participants in the FAS showed no differences among the treatment groups in terms of age group and sex (Table 1). Moreover, all participants were serologically negative at baseline (1:5) against SARS-CoV-2 before the first dosing.

**Table1:** Characteristics of the Participants

| Characteristic | 20-64 years (N=105) |  |  |  | ≥65 years (N=104) |  |  |  |
| --- | --- | --- | --- | --- | --- | --- | --- | --- |
|  | L group (N=30) | M group (N=31) | H group (N=29) | Placebo group (N=15) | L group (N=30) | M group (N=29) | H group (N=31) | Placebo group (N=14) |
| Age, years | 46.3(13.4) | 42.8(12.0) | 44.9(13.9) | 49.6(12.1) | 68.6(3.7) | 70.9(4.3) | 69.7(3.9) | 69.6(3.3) |
| 20-39 years | 11(36.7) | 11(35.5) | 9(31.0) | 5(33.3) | NA | NA | NA | NA |
| 40-64 years | 19(63.3) | 20(64.5) | 20(69.0) | 10(66.7) | NA | NA | NA | NA |
| Sex |  |  |  |  |  |  |  |  |
| Male | 15(50.0) | 15(48.4) | 15(51.7) | 8(53.3) | 15(50.0) | 15(51.7) | 16(51.6) | 7(50.0) |
| Female | 15(50.0) | 16(51.6) | 14(48.3) | 7(46.7) | 15(50.0) | 14(48.3) | 15(48.4) | 7(50.0) |

Data are mean (SD) or n(%).

### Safety

No deaths were reported in this study. One serious adverse event, cerebral infarction, was reported in the placebo group of elderly participants; however, this was judged to be unrelated to the vaccination. In adults, the incidence of adverse reactions from the first dosing to 28 days after the second dosing was 19 of 30 (63.3%), 22 of 31 (71.0%), 22 of 29 (75.9%), and six of 15 (40.0%) in the L, M, H, and placebo groups, respectively. Fever classified as Grade 3 was reported in only one adult in the H group after the second dosing, which resolved the following day.

In elderly participants, the incidence of adverse reactions from the first dosing to 28 days after the second dosing was 14 of 30 (46.7%), 14 of 29 (48.3%), 15 of 31 (48.4%), and six of 15 (40.0%) in the L, M, H, and placebo groups, respectively. No severe (≥ Grade 3) adverse reactions were reported. The most common adverse reaction was injection site pain in both adults and elderly participants (Table 2).

**Table2:** Adverse reactions after the first and the second doses

|  |  | 20-64 years (N=105) |  |  |  | ≥65 years (N=105) |  |  |  |
| --- | --- | --- | --- | --- | --- | --- | --- | --- | --- |
|  |  | L group (N=30) | M group (N=31) | H group (N=29) | Placebo group (N=15) | L group (N=30) | M group (N=29) | H group (N=31) | Placebo group (N=15) |
| Any adverse reaction |  | 19(63.3) | 22(71.0) | 22(75.9) | 6(40.0) | 14(46.7) | 14(48.3) | 15(48.4) | 6(40.0) |
| Solicited Injection site adverse reactions within 0-7 days |  |  |  |  |  |  |  |  |  |
| Erythema | Grade 0 | 3(10.0) | 5(16.1) | 5(17.2) | 1(6.7) | 2(6.7) | 3(10.3) | 2(6.5) | 1(6.7) |
|  | Grade 1 | 0(0.0) | 0(0.0) | 0(0.0) | 0(0.0) | 1(3.3) | 0(0.0) | 0(0.0) | 0(0.0) |
|  | Grade 2 | 0(0.0) | 0(0.0) | 1(3.4) | 0(0.0) | 0(0.0) | 0(0.0) | 0(0.0) | 0(0.0) |
| Swelling | Grade 0 | 1(3.3) | 1(3.2) | 2(6.9) | 0(0.0) | 0(0.0) | 0(0.0) | 0(0.0) | 0(0.0) |
|  | Grade 2 | 0(0.0) | 0(0.0) | 1(3.4) | 0(0.0) | 0(0.0) | 0(0.0) | 0(0.0) | 0(0.0) |
| Induration | Grade 0 | 1(3.3) | 2(6.5) | 3(10.3) | 0(0.0) | 2(6.7) | 0(0.0) | 2(6.5) | 0(0.0) |
| Pain | Grade 1 | 13(43.3) | 16(51.6) | 17(58.6) | 4(26.7) | 9(30.0) | 10(34.5) | 9(29.0) | 3(20.0) |
|  | Grade 2 | 1(3.3) | 0(0.0) | 0(0.0) | 0(0.0) | 0(0.0) | 0(0.0) | 0(0.0) | 0(0.0) |
| Solicited systemic adverse reactions with in 0-7 days |  |  |  |  |  |  |  |  |  |
| Fever | Grade 0 | 0(0.0) | 0(0.0) | 2(6.9) | 1(6.7) | 0(0.0) | 0(0.0) | 0(0.0) | 0(0.0) |
|  | Grade 1 | 0(0.0) | 0(0.0) | 0(0.0) | 0(0.0) | 1(3.3) | 0(0.0) | 0(0.0) | 0(0.0) |
|  | Grade 3 | 0(0.0) | 0(0.0) | 1(3.4) | 0(0.0) | 0(0.0) | 0(0.0) | 0(0.0) | 0(0.0) |
| Headache | Grade 1 | 5(16.7) | 6(19.4) | 4(13.8) | 3(20.0) | 0(0.0) | 0(0.0) | 2(6.5) | 0(0.0) |
|  | Grade 2 | 0(0.0) | 0(0.0) | 2(6.9) | 0(0.0) | 0(0.0) | 0(0.0) | 0(0.0) | 0(0.0) |
| Fatigue | Grade 1 | 4(13.3) | 7(22.6) | 5(17.2) | 2(13.3) | 1(3.3) | 2(6.9) | 2(6.5) | 2(13.3) |
|  | Grade 2 | 1(3.3) | 0(0.0) | 1(3.4) | 0(0.0) | 0(0.0) | 0(0.0) | 0(0.0) | 0(0.0) |
| Nausea | Grade 1 | 0(0.0) | 1(3.2) | 0(0.0) | 0(0.0) | 0(0.0) | 0(0.0) | 0(0.0) | 0(0.0) |
| Muscle pain | Grade 1 | 3(10.0) | 8(25.8) | 6(20.7) | 1(6.7) | 2(6.7) | 3(10.3) | 3(9.7) | 1(6.7) |
|  | Grade 2 | 1(3.3) | 0(0.0) | 0(0.0) | 0(0.0) | 0(0.0) | 0(0.0) | 0(0.0) | 0(0.0) |

Data are n(%), representing the total number of participants who had adverse reactions related to vaccination. The severity of adverse events were graded as Grade0, Grade1 (Mild), Grade2 (Moderate), Grade3 (Severe), and Grade4 (Potentially life threatening).

A dose response in the incidence or severity of adverse reactions was not observed.

The incidence of solicited injection-site adverse events after the third dosing in the H group was 42.9% (12/28) in adults and 25.8% (8/31) in the elderly. The incidence of solicited systemic adverse events was 21.4% (6/28) in adults and 12.9% (4/31) in the elderly. The incidence of adverse reactions after the third dosing was similar to that after the first and second dosing (Table 3).

**Table3:**
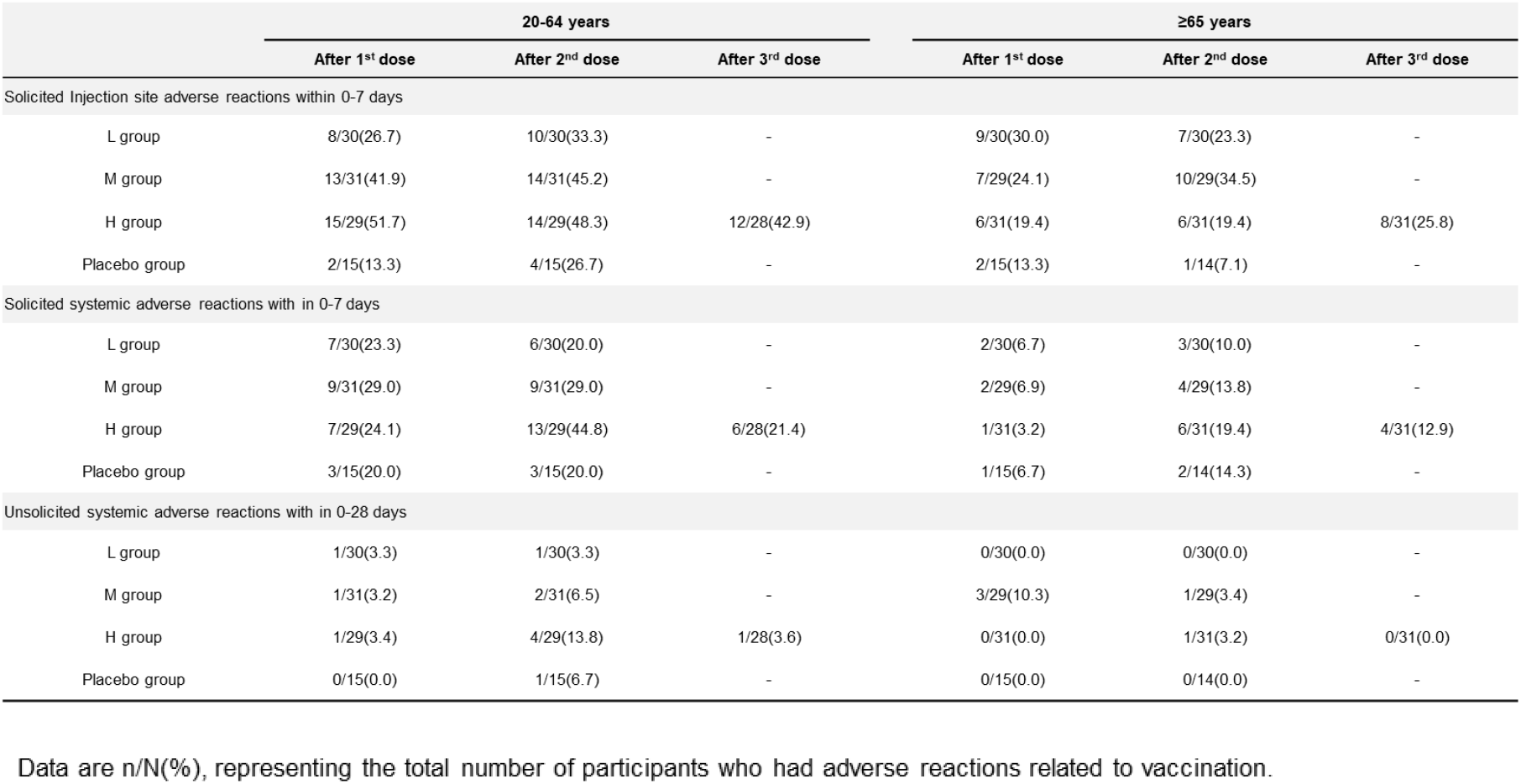
Solicited and unsolicited adverse reactions after the first, the second and third doses

The Th1/Th2 balance, as determined in the exploratory evaluation, showed no particular changes between before and after the first and second dosing, and no differences between ages or groups (Table 4). No changes were observed in cytokine production, and the levels were below the lower limit of quantification for almost all parameters.

**Table4:** Th1 cell / Th2 cell balance after vaccination

| Time Point | 20-64 years |  |  |  | ≥65 years |  |  |  |
| --- | --- | --- | --- | --- | --- | --- | --- | --- |
|  | L group<br>(N=5) | M group<br>(N=7) | H group<br>(N=9) | Placebo group<br>(N=1) | L group<br>(N=6) | M group<br>(N=5) | H group<br>(N=8) | Placebo group<br>(N=3) |
| Before 1<sup>st</sup> dose | 11.08 (5.07) | 18.96 (8.45) | 10.97 (5.38) | 18.90 (NA) | 14.98 (9.15) | 11.94 (6.21) | 15.09 (23.76) | 15.80 (8.60) |
| Day 7 after 1<sup>st</sup> dose | 10.48 (4.39) | 16.03 (6.68) | 9.68 (5.74) | 12.60 (NA) | 12.98 (7.27) | 12.46 (5.59) | 11.74 (13.25) | 14.47 (6.22) |
| Before 2<sup>nd</sup> dose | 10.68 (5.37) | 30.59 (37.75) | 13.16 (8.43) | 17.30 (NA) | 16.03 (9.31) | 14.58 (7.13) | 11.00 (12.19) | 14.17 (7.52) |
| Day 7 after 2<sup>nd</sup> dose | 10.64 (4.70) | 14.99 (5.00) | 10.40 (6.39) | 15.80 (NA) | 13.05 (6.83) | 11.58 (4.56) | 10.53 (13.19) | 10.40 (4.45) |
| Day 28 after 2<sup>nd</sup> dose | 10.20 (4.14) | 15.71 (8.33) | 11.19 (5.64) | 20.10 (NA) | 15.28 (9.55) | 15.74 (9.88) | 13.58 (18.01) | 14.33 (4.44) |

Data are mean (SD). Th1 and Th2 were identified as the number of IFN-γ positive and IL-4-positive cells present, respectively, in the total population of CD4+ T-helper cells.

In addition, none of the participants reported suspected antibody dependent enhancement (ADE) or vaccine-associated enhanced respiratory disease (ERD) from the first dosing to Day 28 after the second vaccination.

### Neutralizing responses

The result of the seroconversion rate (95% CI) for SARS-CoV-2 (D614G) 28 days after the second dosing in each group showed that the seroconversion rate was highest in the H group among both adults and the elderly (adults in the H group: 72.4% [52.8% to 87.3%], elderly in the H group: 45.2% [27.3% to 64.0%]). In addition, neutralizing GMTs (95% CI) 28 days after the second dosing in each group were also highest in the H group (adults in the H group: 12.1 [8.2 to 17.9], elderly in the H group: 7.3 [5.2 to 10.2]) in both adults and the elderly (Table 5).

**Table5:** Seroconversion rate and Geometric mean titers to live SARS-CoV-2 (D614G) induced before and after each dose of KD-414

| Time Point | 20-64 years |  |  |  |  |  | ≥65 years |  |  |  |  |  |
| --- | --- | --- | --- | --- | --- | --- | --- | --- | --- | --- | --- | --- |
|  | L group |  | M group |  | H group |  | L group |  | M group |  | H group |  |
|  | n | Seroconversion /Seropositive rate GMT | n | Seroconversion rate /Seropositive rate GMT | n | Seroconversion rate /Seropositive rate GMT | n | Seroconversion rate /Seropositive rate GMT | n | Seroconversion rate /Seropositive rate GMT | n | Seroconversion rate /Seropositive rate GMT |
| Day 28 after 2<sup>nd</sup> dose | 30 | 36.7%[19.9-56.1]<br>7.1[5.0-10.0] | 31 | 38.7%[21.8-57.8]<br>7.3[4.9-11.0] | 29 | 72.4%[52.8-87.3]<br>12.1[8.2-17.9] | 30 | 33.3%[17.3-52.8]<br>4.8[3.4-6.8] | 29 | 31.0%[15.3-50.8]<br>5.4[3.7-7.8] | 31 | 45.2%[27.3-64.0]<br>7.3[5.2-10.2] |
| Day 91 after 2<sup>nd</sup> dose | 8 | 25.0%[3.2-65.1]<br>3.9[2.3-6.6] | 7 | 28.6%[3.7-71.0]<br>3.7[2.0-6.9] | 29 | 10.3%[2.2-27.4]<br>3.8[2.7-5.5] | 8 | 12.5%[0.3-52.7]<br>3.0[2.0-4.5] | 7 | 0.0%[0.0-34.8]<br>2.8[2.2-3.5] | 31 | 6.5%[0.8-21.4]<br>3.0[2.5-3.6] |
| Before 3<sup>rd</sup> dose | - | - | - | - | 28 | 0%[0.0-10.1]<br>2.6[2.4-2.7] | - | - | - | - | 31 | 0%[0.0-9.2]<br>2.5[NA] |
| Day 28 after 3<sup>rd</sup> dose | - | - | - | - | 28 | 89.3%[71.8-97.7]<br>32.8[21.6-49.9] | - | - | - | - | 31 | 58.1%[39.1-75.5]<br>10.7[6.7-17.0] |
| Day 91 after 3<sup>rd</sup> dose | - | - | - | - | 27 | 70.4%[49.8-86.2]<br>14.0[9.4-20.8] | - | - | - | - | 30 | 43.3%[25.5-62.6]<br>6.8[4.7-9.8] |

Seroconversion/Seropositive rate rates are n/N(%[95%CI]). Geometric mean titers are shown with 95%CI. The ID<sub>50</sub> GMTs were converted to the WHO international standard units (IU/mL) using the conversion factor "0.1458 (for geometric mean)" determined with the WHO International Standard (20/136). Seroconversion was defined as a change from seronegative (ND) at baseline to seropositive (1:10), or four-fold titer increase if the participant was seropositive at baseline (1:5). All participants were serologically negative at baseline (1:5) against SARS-CoV-2 before the first dosing.

**Table6:** Seroconversion rate and geometric mean induced after the second and third doses of KD-414 in H group and immune marker values associated with vaccine effectiveness (VE) against symptomatic infection

|  | Day 28 after 2<sup>nd</sup> dose |  |  |  | Day 28 after 3<sup>rd</sup> dose |  |  |  |
| --- | --- | --- | --- | --- | --- | --- | --- | --- |
|  | n | Seroconversion rate | GMT (ID50) | IU/mL | n | Seroconversion rate | GMT (ID50) | IU/mL |
| 20-30 years | 6 | 66.7(4/6) | 121.6[22.5-657.6] | 17.7[3.3-95.9] | 6 | 100.0(6/6) | 232.8[88.1-615.2] | 33.9[12.8-89.7] |
| 31-40 years | 3 | 66.7(2/3) | 114.8[13.5-978.3] | 16.7[2.0-142.6] | 3 | 100.0(3/3) | 210.7[134.4-330.5] | 30.7[19.6-48.2] |
| 41-50 years | 10 | 40.0(4/10) | 67.2[28.1-160.7] | 9.8[4.1-23.4] | 9 | 66.7(6/9) | 115.9[64.8-207.4] | 16.9[9.4-30.2] |
| 51-60 years | 7 | 42.9(3/7) | 86.7[28.4-264.5] | 12.6[4.1-38.6] | 7 | 42.9(3/7) | 80.9[24.4-268.0] | 11.8[3.6-39.1] |
| 61-70 years | 22 | 36.4(8/22) | 58.1[35.8-94.2] | 8.5[5.2-13.7] | 22 | 50.0(11/22) | 73.8[46.5-117.0] | 10.8[6.8-17.1] |
| ≥71 years | 12 | 16.7(2/12) | 43.9[24.8-77.6] | 6.4[3.6-11.3] | 12 | 25.0(3/12) | 54.0[26.4-110.4] | 7.9[3.8-16.1] |

Seroconversion rates are n/N(%[95%CI]). Geometric mean titers and IU/mL are shown with 95%CI. The ID50 GMTs were converted to the WHO international standard units (IU/mL) using the conversion factor "0.1458 (for geometric mean)" determined with the WHO International Standard (20/136). Seroconversion was defined as a change from seronegative (ND) at baseline to seropositive (80), or four-fold titer increase if the participant was seropositive at baseline (40).

None of the participants in the placebo group showed an increase in neutralizing antibody titers. The seroconversion rate (95% CI) for SARS-CoV-2 (D614G) 28 days after the third dosing in the H group was 89.3% (71.8% to 97.7%) in adults and 58.1% (39.1% to 75.5%) in the elderly, and the GMT was 32.8 (21.6 to 49.9) in adults and 10.7 (6.7 to 17.0) in the elderly. Although the neutralizing antibody titers decreased before the third dosing (5 to 7 months after the second dosing), these were still 2-fold higher after the third dosing compared to that after the second dosing.

Thirteen weeks (91 days) after the second dosing, GMTs were 3.8 (2.7-5.5) in adults and 3.0 (2.5-3.6) in the elderly; however, 13 weeks (91 days) after the third dosing, these were 14.0 (9.4-20.8) in adults and 6.8 (4.7-9.8) in the elderly, showing 3-fold those at 13 weeks after the second dosing. In addition, pseudovirus SARS-CoV-2 (D614) spike protein neutralizing antibody titers in the assay system used in the development of authorized vaccines were measured using serum samples obtained 28 days after the second dosing and 28 days after the third dosing in the H group in the present Phase 1/2 study. The analysis by 10-year age (20-30 years, 31-40 years, 41-50 years, 51-60 years, 61-70 years, ≥71 years) showed that the seroconversion rate (95% CI) against the pseudovirus (D614) was four of six (66.7%), two of three (66.7%), four of 10 (40.0%), two of six (33.3%), nine of 23 (39.1%), and two of 12 (16.7%) 28 days after the second dosing, and six of six (100.0%), three of three (100.0%), six of nine (66.7%), two of six (33.3%), 12 of 23 (52.2%), and three of 12 (25.0%) 28 days after the third dosing, respectively. Moreover, GMTs (95% CI) were 121.6 (22.5-657.6), 114.8 (13.5-978.3), 67.2 (28.1-160.7), 86.7 (28.4-264.5), 58.1 (35.8-94.2), and 43.9 (24.8-77.6) 28 days after the second dosing, and 232.8 (88.1-615.2), 210.7 (134.4-330.5), 115.9 (64.8-207.4), 80.9 (24.4-268.0), 73.8 (46.5-117.0), and 54.0 (26.4-110.4) 28 days after the third dosing, respectively. Furthermore, the IU conversion of pseudovirus (D614) neutralizing GMTs in 10 year segments (20-30 years, 31-40 years, 41-50 years, 51-60 years, 61-70 years, ≥71 years) was 17.7 (3.3-95.9), 16.7 (2.0-142.6), 9.8 (4.1-23.4), 12.6 (4.1-38.6), 8.5 (5.2-13.7), and 6.4 (3.6-11.3) IU/mL, respectively, 28 days after the second dosing and 33.9 (12.8-89.7), 30.7 (19.6-48.2), 16.9 (9.4-30.2), 11.8 (3.6-39.1), 10.8 (6.8-17.1), and 7.9 (3.8-16.1) IU/mL, respectively, 28 days after the third dosing.

## Discussion

Adults aged 20 to 64 years and elderly aged 65 years or older who did not have a history of COVID-19, SARS-CoV-2 infection, or had not received a COVID-19 vaccine, received two intramuscular doses of one of three different dosage formulations of KD-414 (L, M, and H), which showed good tolerability and increased neutralizing antibody titers against SARS-CoV-2 (vaccine strain).

The incidence of adverse reactions was similar among the dose formulations, showing no dose-response relationship. The most common adverse reaction was injection site pain. Most adverse reactions were Grade 1 or Grade 2. For Grade 3 adverse reactions that interfere with activities of daily living, one event of fever, which resolved the following day, was reported after the second dosing in an adult participant in the H group. There were no deaths or related serious adverse events. The incidence of solicited injection-site adverse reactions and fever after two doses of KD-414 were comparable to those of approved inactivated COVID-19 vaccines (BBIBP-CorV, CoronaVac)^13, 14^ that are registered under the WHO Emergency Use Listing Procedure (EUL)^1^ and are used worldwide, but mainly in China. The incidence of injection-site and systemic adverse reactions of KD-414 were relatively lower than those of mRNA vaccines (Comirnaty, Spikevax)^3, 4^, which are currently the main vaccines used for the national vaccination program in Japan.

The incidence of adverse reactions in participants who received the third dose of KD-414 (H) was not particularly different from that after the second dosing, supporting the tolerability of three doses of KD-414. If periodical vaccination is required to prevent a COVID-19 pandemic/endemic in the future, a low incidence of adverse reactions caused by a vaccine would be one of the important factors to promote vaccinees’ intention to active vaccination. There is currently no available COVID-19 vaccine for children under five years of age in Japan. In addition, there are few vaccines that can be safely given to pregnant women. On the other hand, KD-414 is associated with relatively few overall adverse reactions, so it is expected to have potential as a useful vaccine for children and pregnant women.

ADE and ERD, which are related to more severe infectious disease due to viral infection after vaccination, pose a safety risk concern even with COVID-19 vaccines because they have been reported in animal studies of vaccine candidates for other coronaviruses (severe acute respiratory syndrome [SARS] and Middle East respiratory syndrome [MERS])^15, 16^. Th1/Th2 imbalance after SARS-CoV-2 infection is suggested to be one of the possible causes of aggravation^17, 18^. In this study, Th1/Th2 balance and production levels of Th1- and Th2-related cytokines after vaccination with KD-414 were investigated^19^. The results showed that there were no changes in the Th1/Th2 balance and the production levels of related cytokines pre- and post-dose for the first and second doses. Thus, the risk of inducing a Th2-biased Th1/Th2 imbalance with KD-414 vaccination is considered low. Although no suspected ADE/ERD have been confirmed under the current safety monitoring among participants who suffered from COVID-19 after receiving KD-414 after the second or third dosing in this study, further safety monitoring is continuing up to one year after the third dosing.

A dose-response relationship for immunogenicity was confirmed in both adults and elderly participants. Of the three dose formulations, the H group showed the highest immunogenicity 28 days after the second dosing of KD-414.

An age-dependent immune response that induced less neutralizing antibody titers in the elderly compared to other ages was also observed, as with the approved COVID-19 vaccines^20, 21^. The result of stratified analyses by age showed that the neutralizing antibody seroconversion rate in the H group was 100.0% in participants aged ≤ 40 years with high induced neutralizing antibody titers.

Because efficacy of the approved COVID-19 vaccines reportedly diminishes several months after the primary immunization (second dosing), and the third additional (booster) vaccination is given approximately six months after the primary immunization, the safety and immunogenicity of the third dosing of KD-414 (H) were also evaluated in participants of the H group who were continuously participating in this study. The GMT 28 days after the third dosing of KD-414 increased 2-fold of that 28 days after the second dosing. Furthermore, the GMT at 13 weeks after the third dosing tended to be higher than at 13 weeks after the second dosing. Because all of the participants who received the third dose in this study were seronegative for SARS-CoV-2 (D614G) neutralizing antibody titers at about 6 months after the second dosing, an interval shorter than 6 months after the second dosing would be preferable for the third dosing of KD-414.

Currently, there are no established surrogate markers required for the prevention of COVID-19, and neutralizing antibody titers have been measured by various methods in the development of COVID-19 vaccines. Therefore, neutralizing antibody titers of the serum samples obtained in this study were measured by the pseudovirus (D614) neutralizing antibody assay used in clinical studies for Vaxzevria, and were calculated in IU using the conversion factor determined in the assay. Referring to reports on the protective effect (vaccine efficacy [VE]) against Wuhan strains estimated from neutralizing antibody titers^12^, the VE of KD-414 was estimated from pseudovirus (D614) neutralizing antibody titers. The results showed that the titers were 6.4 to 17.7 IU/mL after the second dosing of KD-414, with the VE estimated to be 60% to 70%, and 7.9 to 33.9 IU/mL after the third dosing, with the VE estimated to be 70% to 80%. KD-414 is thus expected to be as effective as currently approved vaccines. In the future, based on the rebalance concept proposed by International Coalition of Medicines Regulatory Authorities (ICMRA)^22^, the efficacy of KD-414 will be confirmed through superiority of immunogenicity to Vaxzevria in the ongoing pivotal Phase 3 study.

A limitation of this study is that we have not yet assessed the effect of KD-414 on induction of both SARS-CoV-2 specific cell-mediated immunity and protective immunity (especially cross-reactivity of neutralizing antibody) against SARS-CoV-2 variants. These assessments will be conducted in ongoing clinical studies. Also, a further clinical safety profile of KD-414 will be evaluated in ongoing pivotal studies.

These findings support progressing KD-414 onto further clinical studies. The recommended dosage was determined to be 10 μg/dose (KD-414 (H)) among the three different doses evaluated in this first-in-human study. KD-414 (H) was well tolerated and safe, and induced high neutralizing antibody responses against SARS-CoV-2, particularly in the age range of 20 to 40 years. In addition, the third dosing of KD-414 (H) showed it to be well tolerated, and it induced a higher and longer-lasting neutralizing antibody response than after the second dosing. Currently, for a three-dose regimen of KD-414 (H) as the primary vaccination, a Phase 2/3 study (jRCT 2071210081), Phase 3 confirmatory study (jRCT 2031210679) and Phase 2/3 study in children aged 6 months to 17 years (jRCT 2031220032) are ongoing.

## Data Availability

All data produced in the present study are available upon reasonable request to the authors.

## ACKNOWLEDGEMENT

This study was funded by Japan Agency for Medical Research and Development (20nk0101603h0001, 21nf0101622h0002).

We thank the institute of Medical Science, the University of Tokyo which provided a virus strain (SARS-CoV-2/UT-HPCo-038/Human/2020/Tokyo).

We also thank all the participants who volunteered for this study.

## AUTHOR CONTRIBUTIONS

MT, KI, and KK designed and coordinated this clinical trial. HN advised technical expertise. KS and ME managed the investigational drug manufacturing and the neutralizing antibody assay. SN and KA managed and analyzed the data of this study. KU contributed to the clinical trial as a medical expert. MT wrote the first draft manuscript, and YS and YO wrote, reviewed and edited. All authors reviewed and approved the final version. All authors had full access to all the data in the study and had final responsibility for the decision to submit for publication.

## CONFLICT OF INTEREST

All authors, except for Kohji Ueda, are employees of KM Biologics Co., Ltd.

Kohji Ueda received a fee from KM Biologics Co., Ltd., for the implementation of this study.

## ETHICS APPROVAL

Hakata Clinic Institutional Review Board gave ethical approval for this study. All necessary participant consent has been obtained before the start of screening.

## Notes

### Clinical Trial

jRCT 2071200106

